# Patterns of Immune Dysregulation in Bipolar Disorder

**DOI:** 10.1101/2024.07.26.24311078

**Authors:** Benney M.R. Argue, Lucas G. Casten, Shaylah McCool, Aysheh Alrfooh, Jenny Gringer Richards, John A. Wemmie, Vincent A. Magnotta, Aislinn J. Williams, Jacob Michaelson, Jess G. Fiedorowicz, Sabrina M. Scroggins, Marie E. Gaine

## Abstract

**Background:** Bipolar disorder is a debilitating mood disorder associated with a high risk of suicide and characterized by immune dysregulation. In this study, we used a multi-faceted approach to better distinguish the pattern of dysregulation of immune profiles in individuals with BD.

**Methods:** We analyzed peripheral blood mononuclear cells (bipolar disorder N=39, control N=30), serum cytokines (bipolar disorder N=86, control N=58), whole blood RNA (bipolar disorder N=25, control N=25), and whole blood DNA (bipolar disorder N=104, control N=66) to identify immune-related differences in participants diagnosed with bipolar disorder compared to controls.

**Results:** Flow cytometry revealed a higher proportion of monocytes in participants with bipolar disorder together with a lower proportion of T helper cells. Additionally, the levels of 18 cytokines were significantly elevated, while two were reduced in participants with bipolar disorder. Most of the cytokines altered in individuals with bipolar disorder were proinflammatory. Forty-nine genes were differentially expressed in our bipolar disorder cohort and further analyses uncovered several immune-related pathways altered in these individuals. Genetic analysis indicated variants associated with inflammatory bowel disease also influences bipolar disorder risk.

**Discussion:** Our findings indicate a significant immune component to bipolar disorder pathophysiology and genetic overlap with inflammatory bowel disease. This comprehensive study supports existing literature, whilst also highlighting novel immune targets altered in individuals with bipolar disorder. Specifically, multiple lines of evidence indicate differences in the peripheral representation of monocytes and T cells are hallmarks of bipolar disorder.

## Introduction

Bipolar Disorder (BD) Type I (BD-I) is a chronic mood disorder, distinguished by recurrent depressive as well as manic episodes, with a lifetime prevalence of just over 1% [1]. Even after receiving treatment, symptom recurrence and further hospitalizations are frequent, prompting the search for better treatment options [2, 3]. Furthermore, individuals with BD are at 20-30 times higher risk of suicide compared to the general population [4]. This indicates a critical need to better understand the pathophysiology of BD including the associated propensity towards suicidal behavior.

The potential value of studying the immune system in BD was first identified when the anti-inflammatory effect of lithium treatment was noted [5]. Numerous differences in immune function have been consistently linked to BD, some of which have been specifically associated with manic and depressive episodes [6]. BD has been associated with elevated monocytes, monocyte activation, and monocyte-derived cytokines [7–10]. Lower and higher T cell activation in individuals with BD has also been observed [11–14]. Meta-analysis of cytokine levels in serum has underlined differences in proinflammatory cytokine and receptor levels including TNF-α [15]. Numerous other cytokines have been associated with BD, but studies conflict on the presence or direction of the changes [16]. Further complicating the interpretation is the incongruence often arising between observations of circulating levels of cells, gene transcripts, and cytokines.

Potentially related to the above observations, several autoimmune diseases have been linked to BD risk, including Crohn’s disease and ulcerative colitis (jointly referred to as Inflammatory Bowel Disease (IBD)) [17–23]. Autoimmune disease was found to be significantly more prevalent in participants with BD compared to controls and those with schizophrenia [24]. Although these findings suggest that immune dysregulation is a facet of BD pathophysiology, identifying the full scope of immune molecules involved and the pathways underlying the dysregulation requires comprehensive evaluation.

The goal of this study was to broadly characterize immune dysregulation in BD. We hypothesized that participants with BD would display higher levels of inflammation and that genetic variants linked to autoimmune disorders would be overrepresented in participants with BD compared to controls. Specifically, we hypothesized that differences mediated by monocytes and T cells would underlie chronically higher inflammation in participants with BD, especially those with a history of suicide attempts. To test our hypothesis, we analyzed immune cell populations, circulating cytokine levels, RNA expression, and DNA variation in individuals with BD and controls.

## Methods and Materials

### Participant Recruitment

Participants were recruited through the Iowa Neuroscience Institute BD Research Program of Excellence after University of Iowa Institutional Review Board approval as previously described [6, 25–27] (**Table 1**). Briefly, individuals between 18 and 70 years old with the capacity to consent and a DSM-5 diagnosis of BD-I were recruited for the group with BD. Prospective controls completed a brief survey and were recruited to balance cases on measures of subjective socioeconomic status (SES), age, race, and sex. Exclusion criteria for both groups included neurological comorbidities, current drug or alcohol abuse, prior loss of consciousness for over ten minutes, or MRI contraindications. Controls were also excluded if they were currently taking medication for depression, schizophrenia, or BD. Participants also completed several assessments as described in Magnotta et al. [27] including the Columbia Suicide Severity Rating Scale. Not every subject was included in every analysis; specific demographics are outlined in **Supplemental Table 1**.

**Table 1.** Participant demographics for samples included in the flow cytometry, cytokine panel, RNA-sequencing, and polygenic propensity experiments.

| Subject Demographics |  | Bipolar Disorder with Suicide Attempt | Bipolar Disorder Without Suicide Attempt | Controls |
| --- | --- | --- | --- | --- |
| Total |  | 55 | 63 | 82 |
| History of suicide attempts |  | 55 | 0 | 1 |
| Number of suicide attempts |  | 3.5 (1-30) | 0 | 1 (1) |
| Body Mass Index (BMI) |  | 30.9 (18.9-50.5) | 30.7 (18.4-59.0) | 27.3 (16.0-44.1) |
| Age |  | 42.0 (18-68) | 38.3 (18-66) | 40.3 (18-70) |
| Mood state | Depressed | 26 | 12 | n/a |
|  | Euthymic | 25 | 48 | n/a |
|  | Manic | 4 | 3 | n/a |
| Ethnicity | Hispanic or Latino | 4 | 3 | 6 |
|  | Not Hispanic or Latino | 51 | 57 | 76 |
|  | Unknown | 0 | 3 | 0 |
| Race | Am. Indian/Alaskan Native | 1 | 1 | 0 |
|  | Asian | 0 | 2 | 3 |
|  | Black/African American | 4 | 2 | 5 |
|  | White | 48 | 57 | 73 |
|  | More than one | 2 | 1 | 0 |
|  | Unknown | 0 | 0 | 1 |
| Sex | Female | 39 | 37 | 55 |
|  | Male | 16 | 26 | 27 |
| Smoker | Unknown | 2 | 0 | 0 |
|  | Never smoker | 18 | 33 | 68 |
|  | Current smoker | 14 | 11 | 5 |
|  | Past smoker | 21 | 19 | 9 |
| Smoking Now | N/A | 3 | 2 | 3 |
|  | Every day | 12 | 9 | 4 |
|  | Some days | 2 | 4 | 1 |
|  | Not at all | 38 | 48 | 74 |

### Flow Cytometry

We first sought to investigate cellular differences related to inflammation in our participants with BD. To do this, we used flow cytometry on fresh samples to quantify the most common immune cell populations. Peripheral blood mononuclear cells (PBMCs) were isolated from 8-10 mL of whole blood collected in EDTA tubes using density gradient separation (Ficoll-Paque PLUS, Cytiva). Samples were then stained for flow cytometric analysis with fluorochrome conjugated antibodies to CD3, CD4, CD14, CD8, and CD19 as previously described [28, 29]. Fluorochrome conjugated, purified mouse immunoglobulin isotype controls were used for background fluorescence. Following staining, cells were fixed in fixation buffer (eBioscience). Flow cytometric data were obtained within 48 hours using either a Becton Dickinson LSR II or a Thermo Fisher Attune NxT. Dead cells were excluded by forward/orthogonal light scatter characteristics. Single cells were identified via FSC-A versus SSC-W. All data was then analyzed using FlowJo software (Tree Star Inc.). Statistical significance was assessed using a two-tailed Student’s t-test, with significance set at α = 0.05.

### Cytokine Panels

To begin to understand the potential functional significance of the cell population differences we observed, we next investigated the serum cytokine levels in individuals with BD. We utilized a protein panel to study 65 immune related proteins simultaneously, which increases the potential for novel findings compared to the single cytokine assays performed previously in BD. To measure cytokine levels in serum, we used the Immune Monitoring 65-Plex Human ProcartaPlex^TM^ Panel (ThermoFisher Scientific), following standard manufacturer protocol. Samples were tested in singlet. To test for batch effect, one sample was replicated across plates. Using two panels and 25 µL of serum from each participant, the cytokine levels of 86 participants with BD and 58 controls were analyzed using a Bio-Plex 1 (BioRad) instrument. Initially, analysis was performed focusing on the impact of suicide attempts in BD, which yielded no significant results in multivariable models. The cytokine data was subsequently incorporated into this paper to provide an additional window into the immune profiles of BD participants. Results were analyzed using non-parametric univariable and multivariable inferential statistics. The Wilcoxon Rank Sum was used to compare BD and control groups for each protein. For multivariable analyses, proteins were rank transformed for linear regression, adjusting for age (continuous, linear), sex (dichotomous), body mass index (BMI; continuous, linear), subjective SES (linear), and current smoking status (dichotomous). Covariates were selected on clinical grounds from an *a priori* statistical analysis plan. We hypothesized that proteins associated with T helper cells will be the most significantly changed including IL-4, IL-10, IL-17a, and TGF-β. To manage Type I error, our primary outcome of interest was IL-17A with other T helper associated proteins as secondary. Molecules whose minimum and maximum concentration in the samples was out of range (OOR) of the Bio-Plex 1 were excluded from the results.

### RNA-sequencing

To investigate immune-related RNA changes associated with BD, we performed mRNA-sequencing using Novogene co. The Paxgene Blood RNA Kit (Qiagen) was used for the initial lysis and processing of the sample, followed by the *mir*Vana miRNA Isolation Kit (Life Technologies) to complete the extraction including a DNase treatment step. 1000 ng of RNA from 25 participants with BD and 25 controls was submitted to Novogene for RNA sequencing. Libraries were pooled and sequenced on the NovaSeq platform using 150 bp paired-end reads. Differential expression of the sequenced libraries was assessed. The edgeR R package (3.22.5) [30, 31] was used to adjust read counts with one scaling normalized factor and then identify differential gene expression between BD and control conditions. Benjamini & Hochberg method was used to adjust p-values to <0.05.

In addition to the gene expression analysis performed by Novogene, we performed enrichment analysis using the clusterProfiler package installed in R (version 4.1.2) [30, 32] and over- representation analysis using the core analyses feature of the Qiagen Ingenuity Pathway Analysis (IPA) software. In the gene set enrichment analysis, all 34,357 genes were included. The most well-known and well-supported gene annotation databases were used: Gene Ontology (GO) and the Kyoto Encyclopedia of Genes and Genomes (KEGG). For the IPA analysis, we included the 2,117 differentially expressed genes (DEGs; p<0.05) identified in BD participants compared to controls.

### Genotyping

We hypothesized that immune-related genetic variation may be associated with BD. To test this hypothesis, we used the PsychArray to identify single nucleotide polymorphisms (SNPs) in the participants. The Gentra Puregene Blood Kit (Qiagen) was used to extract DNA from the blood samples. 15 µL of extracted DNA from each sample was prepared with a concentration of 50-80 ng/µL for submission to the Iowa Institute of Human Genetics, Genomics Division. Samples were genotyped with Illumina’s Infinium PsychArray-24 v1.3 BeadChip (N=188), and genotypes were called using GenomeStudio2.0 software with Illumina’s provided cluster files. PLINK [33] was then used for quality control. Data was filtered down to single nucleotide variation, removing indels and non-biallelic sites. Variants with low minor allele frequency in our sample (< 5%) were removed. All samples had high genotyping rates (>99%). Variants defying Hardy-Weinberg equilibrium were removed (p<1x10^-6^). Four individuals were removed due to relatedness (identity-by-descent cutoff of 0.125), eight were removed based on high heterozygosity rates, and six were removed due to population stratification (did not cluster with the European superpopulation in the 1000 Genomes Project [34]).

A total number of 104 BD and 66 control samples were included in the final analysis. Genotypes for the 170 individuals (350,345 SNPs passing quality control) were imputed to the HRC r1.1 hg19 build reference dataset [35] using Michigan Imputation Server MiniMac4 [36]. Imputation R^2^ scores <0.3 were used to exclude genetic variants and imputed genotype dosages were converted to hard calls using a 90% probability cut-off. A final number of 1,053,095 variants were then subset to HapMap3 variants [37] for polygenic propensity score (PGS) calculations.

### Genetic correlations

Publicly available GWAS summary statistics were obtained for BD [38], IBD [39], Crohn’s disease [39], ulcerative colitis [39], celiac disease [40], type 1 diabetes [41], lupus [42], rheumatoid arthritis [43], and schizophrenia [44]. All sets of GWAS summary statistics underwent basic quality control. Variant positions were converted to hg19 when necessary, and variants with low reported imputation quality scores or allele frequencies were dropped (“info” score <0.9 or minor allele frequency <1%). Linkage disequilibrium score regression (LDSC [45]) was used to compute genetic correlations (r_g_) between BD and immune traits using the 1000 Genomes European LD reference data [34].

### PGS associations

GWAS summary statistics files used in the genetic correlations analysis for BD and schizophrenia were used to compute PGS in the 170 samples with quality-controlled genotype data. PGS were calculated for our cohort using LDpred2 [46] implemented by the “bigsnpr” package [47] in R [30]. A UKBiobank dataset of 362,320 European individuals (provided by the developers of LDpred2) was used to calculate the genetic correlation matrix, estimate heritability, and calculate infinitesimal beta weights. PGS were corrected for population stratification (by regressing out the main effects of the first five genetic principal components).

Associations between BD status and BD PGS was determined with a t-test comparing BD cases and controls. Flow cytometry measures were corrected for age, sex, BMI, and smoking status then z-scaled for association with BD PGS and schizophrenia PGS. Spearman correlations were used to quantify relationships between polygenic propensity for BD or schizophrenia and immune cell population data.

## Results

### BD is associated with altered immune cell populations

The total number of PBMCs did not significantly vary between participants with and without BD (p=0.92), which ensured comparable sample groups. There were significant differences in the percentage of several specific immune cell subtypes within the total PBMCs (**Figure 1**). A lower percentage of CD3+ T cells was found in BD participants (p<0.01; difference between means=11.39±3.90). CD4+ T helper cells specifically were found to comprise a lower percentage of PBMCs in BD participants (p<0.05; difference between means=7.90±3.25). We also observed a significantly greater proportion of CD3- cells in BD participants (p<0.05; difference between means=-8.35±3.94) with the percentage of CD14+ monocytes higher in individuals with BD (p<0.001; difference between means=-7.41±2.06). No significant differences in the percentage of CD8+ T cells and CD19+ B cells were observed (p=0.42 and p=0.24 respectively; **Supplemental Figure 1**). These results suggest an overall disruption in the balance of diverse immune cell populations in BD participants, affecting both innate and adaptive immune cell responses.

**Figure 1.**
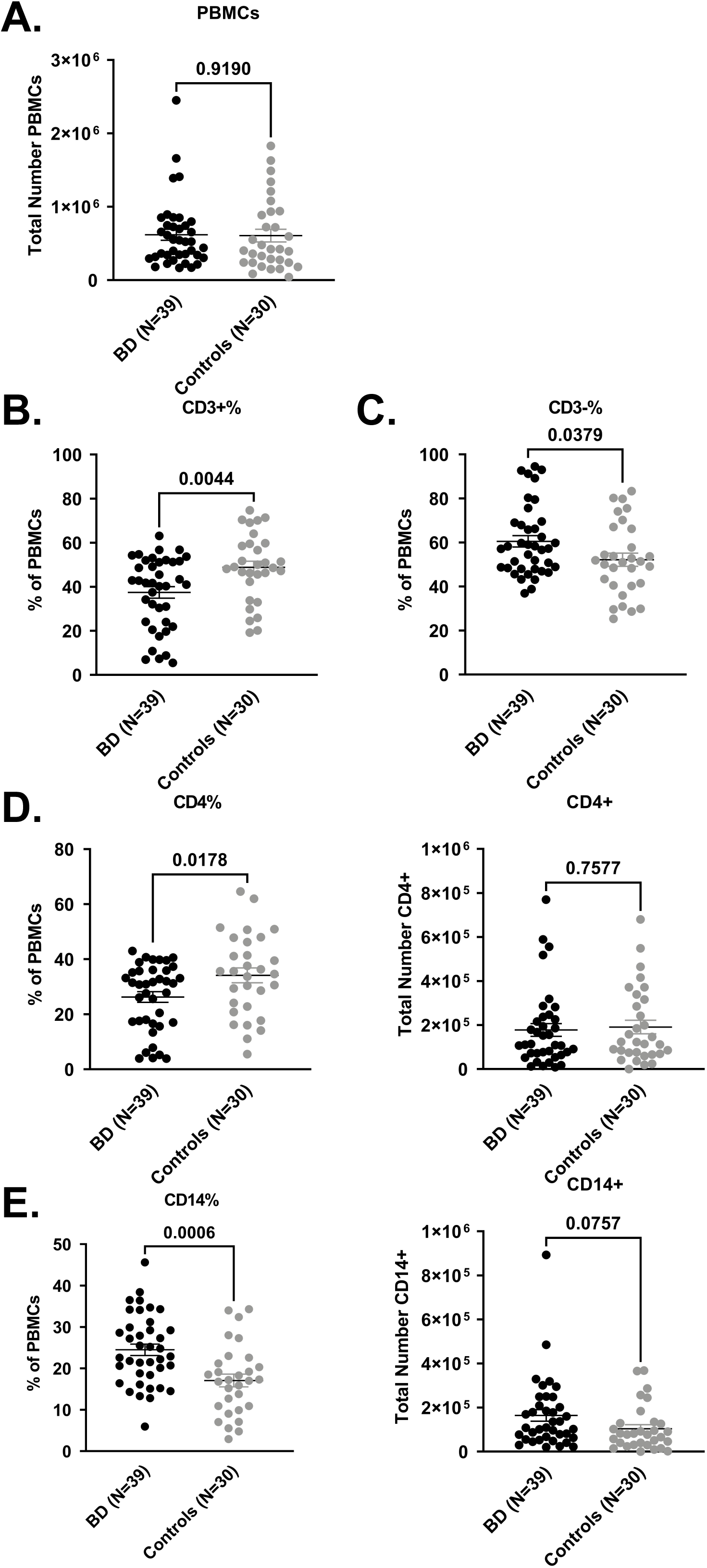
Immune cell population distribution in bipolar disorder (BD) and control participants. Each circle represents one participant, and results are shown with standard error of the mean using an unpaired t-test. **A.** No significant difference in the total PBMCs was found. **B.** In BD participants, we found CD3+ cells constituted a significantly lower percentage of the total PBMCs while (**C.**) a higher percentage of CD3-cells was seen. **D.** A lower proportion of CD4+ cells was observed in participants with BD with no difference in total CD4+ cells. **E.** We also noted a higher percentage of CD14+ cells in BD participants with no difference in total CD14+ cells.

### BD is associated with altered cytokine levels

The concentration of our primary outcome of interest, IL-17A, was not different between BD and controls (p=0.82; **Supplemental Table 2**). However, the expression of 20/65 immune-related molecules were found to be significantly altered (p<0.05) in participants with BD after adjusting for age, sex, BMI, subjective SES, and current smoking status (**Table 2**). Among these were T cell chemoattractants eotaxin and eotaxin-2 (1.51-fold increase in BD, p<0.001; and 1.75-fold increase in BD, p<0.001 respectively). IFNγ levels were also elevated in participants with BD (1.50-fold increase, p<0.001). Cytokines IL-7, IL-12p70, and IL-18 were also elevated in BD participants (1.50-fold increase, p<0.001; 1.33-fold increase, p<0.05; and 1.79-fold increase, p<0.05 respectively). Monocyte-related cytokines MDC, MCP-1, and MCP-2 levels were higher in participants with BD (2.17-fold increase, p<0.001; 2.08-fold increase, p<0.01; and 1.72-fold increase, p<0.01 respectively). We also noted several molecules in the TNF superfamily had higher cytokine concentrations in BD participants: TNFR2 (1.61-fold increase, p<0.001) and TWEAK (1.81-fold increase, p<0.001). MIP-3α and SDF1α levels were also higher in BD participants (3.02-fold increase, p<0.001; and 1.87-fold increase, p<0.001 respectively). Five other cytokines, ENA-78, LIF, HGF, SCF, and CD40L, were all elevated in BD participants (1.57-fold increase, p<0.01; 2.15-fold increase, p<0.01; 1.27-fold increase, p<0.05; 2.15-fold increase, p<0.05; and 1.64-fold increase, p<0.05 respectively). On the other hand, BD participants had significantly lower levels of MIF and IL-16 (0.78-fold change, p<0.001; and 0.34-fold change, p<0.001 respectively).

**Table 2.** Immune molecules differentially expressed in the serum of bipolar disorder participants.

| Molecule | Total Sample<br>N=144 | Subgroup |  | Inferential Statistics |  | Direction<br>of<br>Difference |
| --- | --- | --- | --- | --- | --- | --- |
|  |  | Bipolar Disorder<br>N=86 | Control<br>N=58 | Univariable | Multi-<br>variable |  |
| EOTAXIN | 31.1 (42.2-21.1) | 36.9 (46.6-25.9) | 24.5 (33.5-18.9) | $z=-4.48, p<0.001$ | $p<0.001$ | Higher |
| EOTAXIN-2 | 97.8 (187-61.8) | 128 (831-68.4) | 73.3 (118-53.0) | $z=-4.11, p<0.001$ | $p<0.001$ | Higher |
| IFNγ | 2.8 (3.9-1.9) | 3.3 (4.1-2.4) | 2.2 (3.1-1.4) | $z=-3.85, p<0.001$ | $p<0.001$ | Higher |
| IL-16 | 104 (178-48.8) | 58.8 (116-30.7) | 171 (211-128) | $z=6.31, p<0.001$ | $p<0.001$ | Lower |
| IL-7 | 0.5 (0.9-0.3) | 0.6 (1.2-0.3) | 0.4 (0.6-0.2) | $z=-3.71, p<0.001$ | $p<0.001$ | Higher |
| MDC | 47.0 (136-4.3) | 71.1 (252-13.4) | 32.7 (53.5-<OOR) | $z=-3.64, p<0.001$ | $p<0.001$ | Higher |
| MIP-3α | 17.7 (40.0-6.3) | 30.2 (51.9-9.9) | 10.0 (17.7-4.8) | $z=-4.93, p<0.001$ | $p<0.001$ | Higher |
| SDF-1α | 625 (1481-476) | 967 (1818-556) | 516 (624-449) | $z=-5.15, p<0.001$ | $p<0.001$ | Higher |
| TNFR2 | 49.0 (81.0-35.7) | 64.6 (106.4-40.3) | 40.1 (51.3-33.9) | $z=-4.49, p<0.001$ | $p<0.001$ | Higher |
| TWEAK | 622 (1139-389) | 858 (1457-437) | 473 (745-358) | $z=-3.64, p<0.001$ | $p<0.001$ | Higher |
| MIF | 18.1 (23.3-12.8) | 15.5 (21.6-9.6) | 19.8 (24.5-16.7) | $z=3.28, p=0.001$ | $p<0.001$ | Lower |
| MCP-1 | 19.9 (48.3-8.3) | 28.7 (68.2-10.4) | 13.8 (27.0-<OOR) | $z=-3.46, p<0.001$ | $p=0.001$ | Higher |
| ENA-78 | 72.7 (136-43.8) | 92.6 (171-53.3) | 59.1 (89.0-36.9) | $z=-3.03, p=0.002$ | $p=0.006$ | Higher |
| MCP-2 | 4.1 (9.0-2.4) | 5.5 (11.6-2.5) | 3.2 (5.8-2.1) | $z=-2.42, p=0.02$ | $p=0.007$ | Higher |
| LIF | 2.8 (7.0-1.2) | 3.3 (5.8-2.0) | 1.5 (9.1-<OOR) | $z=-2.17, p=0.03$ | $p=0.008$ | Higher |
| HGF | 31.5 (62.0-19.2) | 36.8 (104-21.9) | 29.0 (42.4-14.4) | $z=-2.70, p=0.007$ | $p=0.02$ | Higher |
| IL-12p70 | 1.0 (1.4-0.8) | 1.2 (1.5-0.9) | 0.9 (1.2-0.7) | $z=-2.31, p=0.02$ | $p=0.02$ | Higher |
| IL-18 | 3.3 (9.9-0.3) | 4.3 (14.6-<OOR) | 2.4 (4.7-0.4) | $z=-2.04, p=0.04$ | $p=0.03$ | Higher |
| SCF | 4.2 (8.2-0.8) | 5.6 (9.6-1.7) | 2.6 (6.2-0.4) | $z=-2.46, p=0.01$ | $p=0.03$ | Higher |
| CD40L | 24.3 (75.0-4.2) | 26.4 (73.8-8.1) | 16.1 (109.2-<OOR) | $z=-1.23, p=0.22$ | $p=0.03$ | Higher |

When considering suicide-specific changes in the subsample with BD (N=38), the analysis of cytokine levels associated with a history of suicide attempts in BD yielded one notable result. Lower CD40L was associated with a history of suicide attempts in BD participants (0.64-fold change, p<0.05), however this finding did not remain significant following multivariable testing (p=0.15; **Supplemental Table 2**).

### BD is associated with altered immune gene expression levels

RNA-sequencing revealed 49 DEGs in BD participants (adjusted p<0.05; **Supplemental Table 3**), with 44 up-regulated and 5 down-regulated. *IFI27*, which is involved in IFNγ signaling, was significantly higher in participants with BD (log2 fold change=1.79). Genes implicated in IL-7 signaling were also upregulated, specifically *BCL2L1* and *ABALON* (log2 fold change=1.22; and log2 fold change=1.18 respectively). We noted T-reg enriched *NEDD4L* displayed higher expression levels in individuals with BD (log2 fold change=0.76). All significant immune gene findings are reported in **Supplemental Table 3**. Using our generated gene lists, we performed integrative functional bioinformatic analyses to identify pathways enriched for DEGs and identified several immune-related pathways (**Supplemental Tables 4** and **5**) including innate immune response (adjusted p<0.01).

### BD, but not schizophrenia, genetic propensity is associated with immune cell populations

To identify whether genetic risk loci related to psychiatric disorders was associated with components in our data, we use BD and schizophrenia PGS. As a positive control, we found that individuals with BD carry significantly higher BD PGS (t=5.34, p=4.00x10^-7^; **Figure 2A**). Next, we tested if BD genetic propensity was associated with immune cell populations. Flow cytometry results were z-scored after correcting for age, sex, BMI, and current smoking status. These corrected immune cell population scores were correlated with PGS for BD, showing a significant positive relationship between polygenic propensity for BD and percentage of CD14+ cells (Pearson’s R=0.27, p<0.05; **Figure 2B**). Notably, using a schizophrenia PGS showed no correlation (Pearson’s R=0.08, p=0.53; **Supplemental Figure 2**), suggesting that this correlation is specific to BD as opposed to psychiatric disorder risk in general. These results support our flow cytometry finding that a higher proportion of monocytes are present in individuals with BD and expound on this observation by identifying a linear relationship between genetic propensity for BD and higher monocyte percentage.

**Figure 2.**
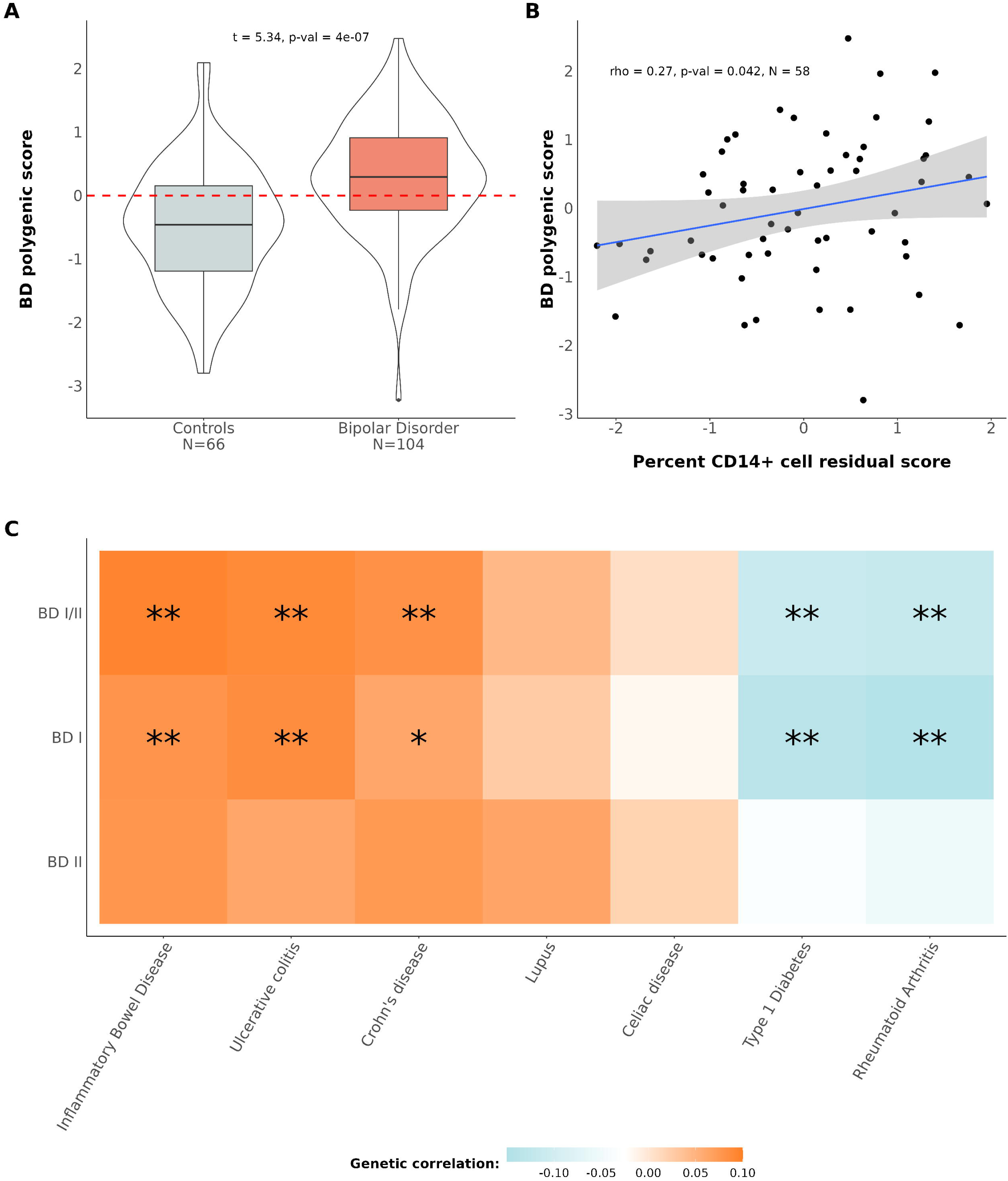
Polygenic propensity score analyses. **A.** Bipolar disorder (BD) polygenic risk is higher in BD participants as shown by Z-scores compared between control and BD participants. **B.** BD PGS is associated with CD14%. Each circle represents data from one participant included in both the genetic and cellular experiments. **C.** Linkage disequilibrium score regression (LDSC) genetic correlations (r_g_) between BD and immune diseases show specific overlap. Double asterisks (“**”) denote associations with a FDR adjusted p-value <0.05. Single asterisks (“*”) denote associations with an unadjusted p<0.05.

### Specific autoimmune diseases share genetic risk with BD-I

To identify whether genetic risk loci are shared between immune disorders and BD we conducted a genetic correlation analysis using LDSC in the UKBiobank dataset [45]. Because of the larger dataset, we were able to investigate BD-I and BD Type II (BD-II) separately. We replicated findings of comparable analyses performed with older GWAS summary statistics [48], finding mixed genetic correlation effects between immune disorders and BD (**Figure 2C**). A significant positive correlation between BD-I and IBD (r_g_=0.09, FDR adjusted p<0.05), ulcerative colitis (r_g_=0.08, FDR adjusted p<0.05), and Crohn’s disease (r_g_=0.06, unadjusted p<0.05, FDR adjusted p=0.07) was found. Significant negative correlations between BD-I and immune disorders, included: Type 1 diabetes (r_g_=-0.13, FDR adjusted p<0.001) and rheumatoid arthritis (r_g_=-0.14, FDR adjusted p<0.001). These results suggest that there is specificity in which immune disorders share genetic risk loci with BD-I, and genetic risk factors for IBD increase risk for BD-I. Despite similar findings when combining BD subtypes, no correlation was found with BD-II.

We attempted to validate the genetic correlation results in our own dataset. We computed immune PGS for our sample and tested for differences between BD and controls. None of the immune PGS were significantly different between BD and controls in our dataset, possibly due to the much smaller sample size (data not shown).

## Discussion

In this analysis of immune dysregulation in BD we evaluated live cellular, serum cytokine, gene expression, and genetic variation, and identified widespread immunological differences associated with BD. Our strongest findings, derived from multiple experimental approaches, indicate that lower T helper cell proportions and higher monocyte populations may be involved in the pathophysiology of BD.

The lower percentage of T helper cells seen in participants with BD supports findings linking susceptibility to develop BD to lower percentages of T cells [13]. However, it conflicts with previous reports which found no difference or higher percentages of T cells [11, 14]. Upstream of T cells, we found IL-16 was lower in participants with BD. IL-16 activates and primes T cells, particularly T helper 1 (Th1) cells [49]. Despite the overall decreased proportion of T helper cells, we noted several associated circulating factors of these cells were higher in BD. These data are in alignment with the widely accepted concept that immune dysregulation is not solely due to alterations in cell numbers, but also involves alterations in cellular function. We found that IL-7 concentration, which is protective of T cells, was higher in participants with BD. Elevated IL-7 has been previously associated with BD offspring who go on to develop a mood disorder [50]. Corresponding evidence of higher IL-7 was identified through gene expression analysis. *BCL2L1* and *ABALON*, an antisense long noncoding RNA in *BCL2L1*, were both significantly elevated in BD participants. *BCL2L1* is an antiapoptotic gene encoding the protein Bcl-xL, which is upregulated by IL-7 [51]. IFNγ, CD40L, LIF, and MIF were also elevated in BD participants, indicating T cell activation [52–55]. IP-10 and MIP-3α, which are chemotactic for T cells, were also higher in participants with BD, further implicating T cells in the pathophysiology [56, 57]. Eotaxin and eotaxin2, both of which attract Th2 cells to sites of inflammation, were higher in participants with BD, consistent with other studies [7, 58, 59]. Likewise, we observed elevated HGF levels, which have been associated with higher Th2 responses [60]. Overall, we observed a decreased proportion of T helper cells, however our RNA and cytokine analyses indicate higher Th1 and Th2 activation. Further studies are needed to more accurately categorize alterations in both the specific populations and functions of circulating T helper cells in individuals with BD.

The higher proportion of monocytes in individuals with BD aligns with the findings from our cytokine panel that included many key factors (MCP-1, MCP-2, MIP-3α, IP-10, HGF, and MDC) related to monocyte activation, migration, and infiltration [61–63]. The monocyte alterations seen reflects other BD studies showing altered cytokine levels [7, 64], higher monocyte proportion [8], increased monocyte activation, and the induction of proinflammatory cytokine release in cultured human monocytes [9, 10]. Despite some conflicting results [65], our findings are supported by others that found elevated gene expression and cytokine concentrations of MCP- 1, HGF, TWEAK, and MIP-3α in individuals with BD [10, 58, 65–68]. Notably, genetic variants of MCP-1 have also been identified as a risk factor for BD [69, 70], suggesting an immune-related genetic predisposition in BD. This complements our genetic analysis, which correlated higher BD PGS with higher percentage of monocytes (CD14+%). This positive linear correlation between genetic propensity for BD and monocyte levels strengthens the hypothesis that increased monocyte activation is a key factor in the development of BD.

Our genetic analysis also identified a positive correlation between IBD and BD, suggesting shared underlying mechanisms. Prior observational and genetic research supports our finding [17–23], while other studies have found no association [71–73]. Other lines of evidence from our studies also suggest a link with IBD. For several T cell-related cytokines that we saw were elevated in BD, similar associations have been made with IBD, including higher eotaxin in ulcerative colitis, elevated IL-7 and IFNγ in Crohn’s disease, and higher IL-18 in both conditions [74, 75]. Additionally, our findings of higher monocytes and monocyte-related proteins including MCP-1 and TWEAK have also been seen in IBD [76–79]. Notably, TWEAK is a protein mainly secreted by monocytes/macrophages that induces the production of MCP-1 [80], but it has also been linked to intestinal dysfunction [81] and neuroinflammation [82, 83], suggesting a potential role in both intestinal and brain dysregulation. Barrier disruption, of the intestines in IBD and the blood-brain barrier in BD [84], is a notable parallel. Several inflammatory markers upregulated in BD and IBD, including TWEAK and MCP-1, have been found to damage both of these barriers [85, 86], suggesting one mechanism by which these overlapping immune responses may influence both pathologies. Neuroinflammation has previously been associated with BD and several inflammatory markers noted in our study have been linked to neuroinflammation [64, 87–92]. Although it cannot be presumed that circulating levels will reflect levels in the brain, taken together, these studies suggest that our findings may represent a circulating phenotype for neuroinflammation. Overall, the dysfunction in T helper cells and monocytes could explain, at least in part, the mechanisms shared by IBD and BD. Alternatively, mood symptoms may activate evolutionarily conserved pathways by which stress activates the sympathetic nervous system to promote innate immunity and proinflammatory cytokine production [93–96].

Our results highlight a widespread cascade of primarily proinflammatory immune changes that occur in BD; however, we are unable to pinpoint the underlying instigating factors. To probe the causality of the immune dysregulation we have observed, innovative longitudinal study design is likely required. There are other limitations to our study that should be noted. We did not assess the mood cycle of participants at the time of sample acquisition. Prior studies have found inflammatory patterns associated with either manic or depressive phases of the disorder [6], but our results do not clarify which immune differences may be cycle specific. We also did not have access to information regarding additional inflammatory symptoms that may have altered immune patterns in specific participants. Finally, pharmacologic factors were not assessed, which is important to note as some medications commonly prescribed to individuals with BD, such as lithium, can impact the immune system [9, 10]. Regarding the genetic analyses, we limited our data to Europeans and therefore these findings may not generalize to other populations. These study components may also explain some of the conflicting reports present in the field.

## Conclusion

Overall, our results have identified multifaceted immune system alterations in individuals with BD. The majority of the gene expression and cytokine differences were proinflammatory in nature, indicating that a global increase in inflammation is a characteristic of BD. In addition, we identified significant molecular overlap between IBD and BD. We propose that the immune dysfunction observed in BD, which resembles that seen in IBD, may lead to brain infiltration through a compromised blood-brain barrier, resulting in neuroinflammation. Because this genetic signature is present before clinical onset, it would suggest that it is a causal factor in BD, albeit in the presence of additional risk factors. However, it is also possible that the altered immune response associated with BD is a consequence of the disorder rather than a cause through the sympathetic nervous system. A detailed understanding of the immune system in individuals with BD is needed in to treat immune-related comorbidities of BD, and BD itself. Furthermore, a step towards reducing heterogeneity in the BD population may include immune profiles as biomarkers and/or therapeutic targets. Our findings highlight the importance of studying immune function in BD, and the need for more work in this area.

## Data Availability

All data produced in the present study are available upon reasonable request to the authors

## Acknowledgements

The authors would like to thank the study participants for their willingness to participate in the study. We would also like to thank Hsiang Wen for sample collection and management. Parts of the data presented herein were obtained at the Flow Cytometry Facility, which is a Carver College of Medicine / Holden Comprehensive Cancer Center core research facility at the University of Iowa. The facility is funded through user fees and the generous financial support of the Carver College of Medicine, Holden Comprehensive Cancer Center, and Iowa City Veteran’s Administration Medical Center. Additional data were obtained at the Genomics Division of the Iowa Institute of Human Genetics which is supported, in part, by the University of Iowa Carver College of Medicine.

## Grant Support

This study was supported by an Iowa Neuroscience Institute Research Program of Excellence grant with philanthropy from the Roy J. Carver Charitable Trust (JAW), a US Department of Veterans Affairs Merit Review Award (JAW) and a US Department of Veterans Affairs Senior Clinician Scientist (JAW). It was also supported by an EHSRC Career Enhancement award and a EHSRC Pilot grant (NIH P30 ES005605; MEG) and by the National Center for Advancing Translational Sciences of the National Institutes of Health (UL1TR002537) and the National Institute of Mental Health (NIMH R01MH125838 and R01MH111578 to VAM and JAW). Funding for this work was given to SS through the OB/GYN department at the University of Iowa Hospitals & Clinics, as well as the AHA with grant 19IPLOI34760288, and the NIH with grants NCATS 3UL1TR002537-03W1, NHLBI 1KO1HL155240-01, and NCATS UL1TR002494. Additional funding for this work came from the National Institutes of Health through a Predoctoral Training Grant (T32GM008629 to LGC).

## Disclosures

The authors have no conflicts to declare.

## Supplemental materials

- Supplemental Table 1: Subject demographics in the four experiments
- Supplemental Table 2: Complete cytokine panel results including suicide analysis
- Supplemental Table 3: All significant RNA-sequencing results
- Supplemental Table 4: All significantly enriched GO pathways
- Supplemental Table 5: All significantly enriched IPA canonical pathways
- Supplemental Figure 1: Non-significant immune cell population distribution results in BD.
- Supplemental Figure 2: Schizophrenia PGS is not associated with CD14%.

---

**Supplemental Figure 1:** Non-significant immune cell population distribution results in bipolar disorder (BD). Each circle represents one participant, and results are shown with standard error of the mean using an unpaired t-test. We found no significant difference in percentage or total numbers of **A.** CD8+ or **B.** CD19+ cells.

**Supplemental Figure 2.** Schizophrenia polygenic propensity score is not associated with CD14%. Each circle represents data from one participant included in both the genetic and cellular experiments.

## Notes

### Competing Interest Statement

The authors have declared no competing interest.

### Author Declarations

The Institutional Review Board of the University of Iowa gave ethical approval for this work.

