## Supplementary figures and images for "Patterns of Immune Dysregulation in Bipolar Disorder"

### Supplemental Figure 1

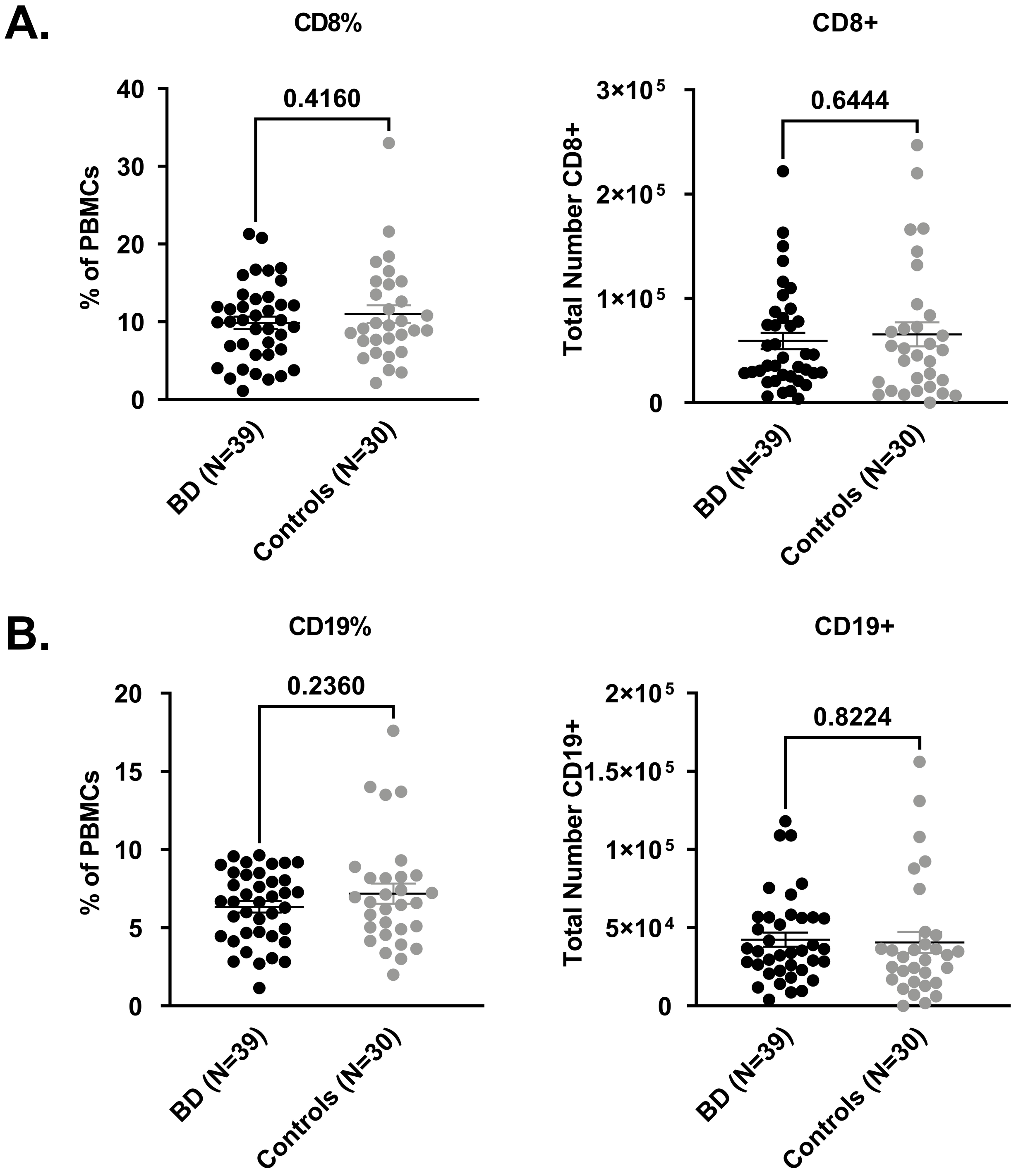

### Supplemental Figure 2

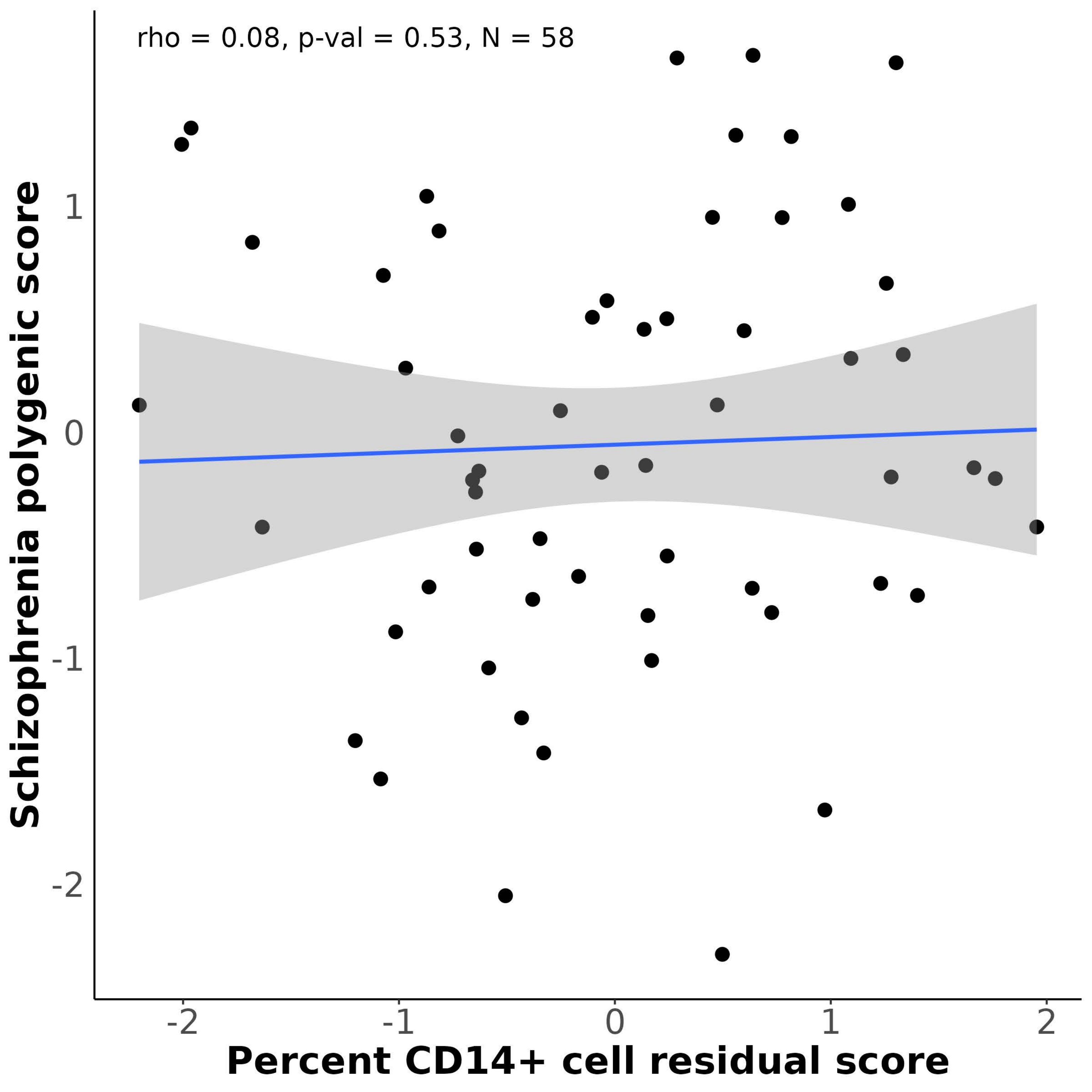
